# How a Positive COVID-19 diagnosis affects the physical, social and psychological wellbeing of people in the United Arab Emirates? An Explorative Qualitative Study

**DOI:** 10.1101/2021.09.28.21264265

**Authors:** Mouza AlKuwaiti, Bayan Abu Hamada, Noof AlJeneibi, Marília Silva Paulo, Iffat Elbarazi

## Abstract

**Purpose:** Exploring the effect of COVID-19 diagnosis on the individual has not been explored through an exploratory qualitative approach. This study aims to explore the physical, social, and psychological impact of the diagnosis on the individual through online interviews.

**Method:** A qualitative study approach using online interviews was conducted. A sample of 30 participants of different age groups, gender, and nationalities were interviewed to explore the impact of a positive COVID-19 diagnosis on their physical, mental, social, psychological health, and lifestyle practices. An interview guide was created based on coping strategy model and conceptual framework of coping strategies. All interviews were recorded then transcribed after obtaining written consent from participants. Ethics approval was obtained from the United Arab Emirates Social Science Ethics Committee. NVIVO software was used for thematic analysis based on both identified coping models to highlight the most important feelings and emotions, family support, and changes in lifestyle that may impact the COVID-19 patient and family. Researchers identified the themes separately and then verified themes in one meeting.

**Results:** Major themes include the physical effects, social effects, psychological effects, spiritual effects, and lifestyle effects. Emerging themes include coping mechanisms, trust in authorities and health care system, appreciation of the role of the government, conspiracy theories, and media roles. Those who had a positive infection towards the end of 2020 and in 2021 described having fewer negative emotions and better psychological resilience.

**Conclusion:** The findings of this study indicate that people diagnosed with COVID-19 have perceived a very good support in terms of their physical health from the government and health authorities, but require social, psychological, and educational support during the infection period and post-recovery.

## Background

People around the world are testing positive for SARS-CoV-2, after one year and a half of the beginning of the pandemic around 300 to 400,000 are being diagnosed daily (Worldmeter.com 2021). The world population is seeing unprecedented changes in their daily lives that can affect their occupation, income, and their health. These challenges by themselves can be stressful, overwhelming, and cause strong emotions in adults and children (Arora et al. 2020; Johal 2009; Stamu-O’Brien et al. 2020; Vindegaard and Benros 2020). On top of these stressors, when a person realizes the diagnose of COVID-19 the increment of all this stress and mixed emotions is believed to be exponential for the person itself and the family members and friends. Viruses being unseen, cause psychological distress including fear, denial, and anxiety in the general population (Pappas et al. 2009). A study conducted in Japan demonstrated that close contacts with COVID-19 patients (family and friends) suffer from high psychological distress suggesting that the establishment and implementation of mental health and psychosocial support measures tailored to family and close relatives and friends of COVID-19 patients are warranted (Tanoue et al. 2020).

During the past year, so many studies focused on the mental health of the general population and the effect of the pandemic on communities and certain groups (Ghosh et al. 2020; Guo et al. 2020; Hossain et al. 2020; Pimenta et al. 2020; Semo and Frissa 2020; Stip, Mugaddam, and Amiri 2020). Studies linked the changes of lifestyle including social isolation, online learning, and strict lockdowns on people’s mental, social and physical health (Munasinghe et al. 2020; SK et al. 2020). A recent systematic review found that two studies only conducted on COVID-19 patients found a high level of post-traumatic stress symptoms (PTSS) and a significantly higher level of depressive symptoms. Patients with preexisting psychiatric disorders reported worsening of psychiatric symptoms (Vindegaard and Benros 2020). Among the reported psychological distress caused by COVID-19, there is loneliness; anxiety, panic, PTSS, stigma, obsessive behaviors, burnout, hoarding, paranoia, substance abuse and depression, outbursts of racism, stigmatization, and xenophobia against particular communities are also being widely reported.

A qualitative study of the psychological experience of COVID-19 patients during hospitalization conducted in China discussed five main areas affecting patients with COVID-19: attitudes toward the disease (fear, denial, and stigma); stress due to the nature of the disease, quarantine measures, and concerns regarding the health of family members; physical and emotional responses, like lifestyle changes in diet, sleep, and behavior; supportive factors included psychological adjustments, medical care, and family and social support: and finally, gratitude through the cherishing of life, family, bravery, and tenacity (Sun et al. 2021). Many studies have explored the effect of the pandemic on the mental health of general and specific populations. However, so far exploring the effect and the impact of the COVID-19 diagnosis on the individual and the experience of the infected person has not been explored through an exploratory qualitative approach in the United Arab Emirates (UAE).

The UAE is considered a relatively young country (50 years old) that emerged from the federation of seven emirates. The country has one of the most competitive health systems in response to its unique population (Paulo, Loney, and Lapão 2017). The population is considered unique, although similar to other neighboring countries, as from the 9.7 million people (2019), 87% were expatriates and there is a bigger proportion of males (69%) compared to females (31%) (Dubai Online n.d.). Another characteristic of the UAE population is the relatively young population, where 84.3% are in the age group of 15 to 64 years old (Dubai Statistics Center 2020; Statistics Center of Abu Dhabi 2019; World Bank 2020).

To provide an overview of the mental physical and social effects and complications of COVID-19 infection among patients in the UAE it is highly important to explore patients’ experiences through a qualitative approach, which allows us to best understand patients’ feelings, experiences, and differences. The conceptual framework of coping and coping strategies framework will be used as the foundation for our interview process. This study intends to contribute significantly with complementary data for the quantitative research, helping policymakers to understand the patients’ needs and complimenting the impressive epidemiologic research that has been done in the country on this specific topic.

To explore the physical, mental, and psychosocial impact of a positive COVID-19 on UAE residents.

## Method

This study is being reported according to the COnsolidated criteria for REporting Qualitative research) (COREQ) (Tong, Sainsbury, and Craig 2007).

The study used a qualitative approach using online face-to-face interviews. Snowballing and convenience sampling was used to identify residents who have had a COVID-19 infection during February 2020 and April 2021. A Sample of 28 participants, 25% males and 75% females, of different age groups (23-60) and different nationalities (50% Emiratis) were interviewed. An interview guide was created based on (Lazzarus and Folkman 1984) (Lazzarus and Folkman 1984) Coping strategy model and (Burr & Klein 1994) conceptual framework of coping strategies.

## Research team and reflexivity

### Personal characteristics

#### Interviewer characteristics

IE, an experienced qualitative researcher and the principal investigator (PI), trained two research assistants (BH and MAL) in conducting interviews and in identifying themes. One research assistant holds a master’s in public health (MPH) and another is a MPH student and a MBSS graduate, the PI holds a DPH in qualitative research methods. The PI has supervised the entire process of the interviews.

### Relationship with participants

#### Relationship established

Participants were recruited randomly from the community after we advertise through WhatsApp that we are recruiting. After few people responded we started asking them to identify and recommend other participants from their circle.

#### Participant knowledge of the interviewer

Few of the participants were known to at least one of the researchers. However, all participants received assurance of confidentiality and privacy. We made sure to assign the known participants to one of the researchers who do not know have acquaintance with.

#### Participants characteristics and sample size

Participants were UAE residents, who have had a positive COVID-19 diagnosis and who have recovered between the period of February 2020 and April 2021. Around 50 participants were invited only 28 agreed to participate. We tried to have a representation of both genders, different nationalities, and age groups.

### Study design

#### Theoretical framework

The study used a qualitative inductive approach using online face-to-face interviews through zoom or MS teams based on the participants’ preference.

An interview guide was created based on (Lazzarus and Folkman 1984) Coping strategy model and (Burr & Klein 1994) conceptual framework of coping strategies. The main PI developed the interview guide. The other research team members reviewed and piloted the interview guide. One of the researchers has had COVID-19 herself and has a background in psychology (NAJ). She reviewed the interview guide and added questions that were related to social impact.

### Participant selection

#### Sampling

Snowballing and convenience sampling was used to identify residents who have had a COVID-19 infection. Participants were identified through WhatsApp via contacts and then via the snowball technique.

#### Method approach

Participants were approached by phone and via WhatsApp. Only those who agreed to participate through zoom or teams were provided a written consent form were included in the study. All interviews were audio-recorded and then transcribed. Most of the interviews were conducted in Arabic then transcribed and translated. NVIVO qualitative analysis software was used to help in identifying and organizing themes.

### Setting

All interviews were conducted online due to the COVID-19 restrictions. Ethics approval was obtained UAEU Social Science Ethics Committee Approval number: ERS_2021_7264.

### Data collection

#### Interview guide

The interview guide was developed in both Arabic and English language with questions exploring the mental, social, psychological, and lifestyle impact of a positive COVID-19. The interview guide was reviewed by the research team who agreed on the final questions and concepts to be addressed through interviews. Piloting for the interview guide was conducted with five participants working in the health care field and are included within the main sample reported.

All participants received full information about the study aims goals, the possibility to withdraw from the research at any time, and confidentiality. Interviews were conducted by trained researchers in qualitative interviews in both Arabic and English based on the participants’ preferences.

#### Repeated interviews

All interviews were conducted once and are reported

#### Audio/visual recording

All interviews were recorded then transcribed after obtaining written consent from participants. A research assistant collected all recordings, transcribed them, and translated them from Arabic to English. All interviews were imported to NVivo 12 for analysis and coding.

#### Duration

Each interview lasted between 40-60 minutes.

#### Data saturation

Data saturation was reached after interviewing 15 participants however we decided to keep interviewing participants to ensure we have a wide variety and more representation of the UAE community.

#### Transcript returned

To ensure researchers’ reflexivity and to reduce bias some of the interviews were transcribed and returned to few participants for review and approval. Participants showed no concerns and were happy with transcribed words.

## Analysis and findings

### Data analysis

#### Number of data coders

Framework thematic analysis was performed. A Thematic tree was created to highlight the most common themes (Figure 1). IE and MSP have revised the transcripts of the interview for triangulation. The research team was given training by the PI on how to extract themes. After all, researchers extracted their themes. IE, MA, MSA agreed on final themes.

**Fig 1.**
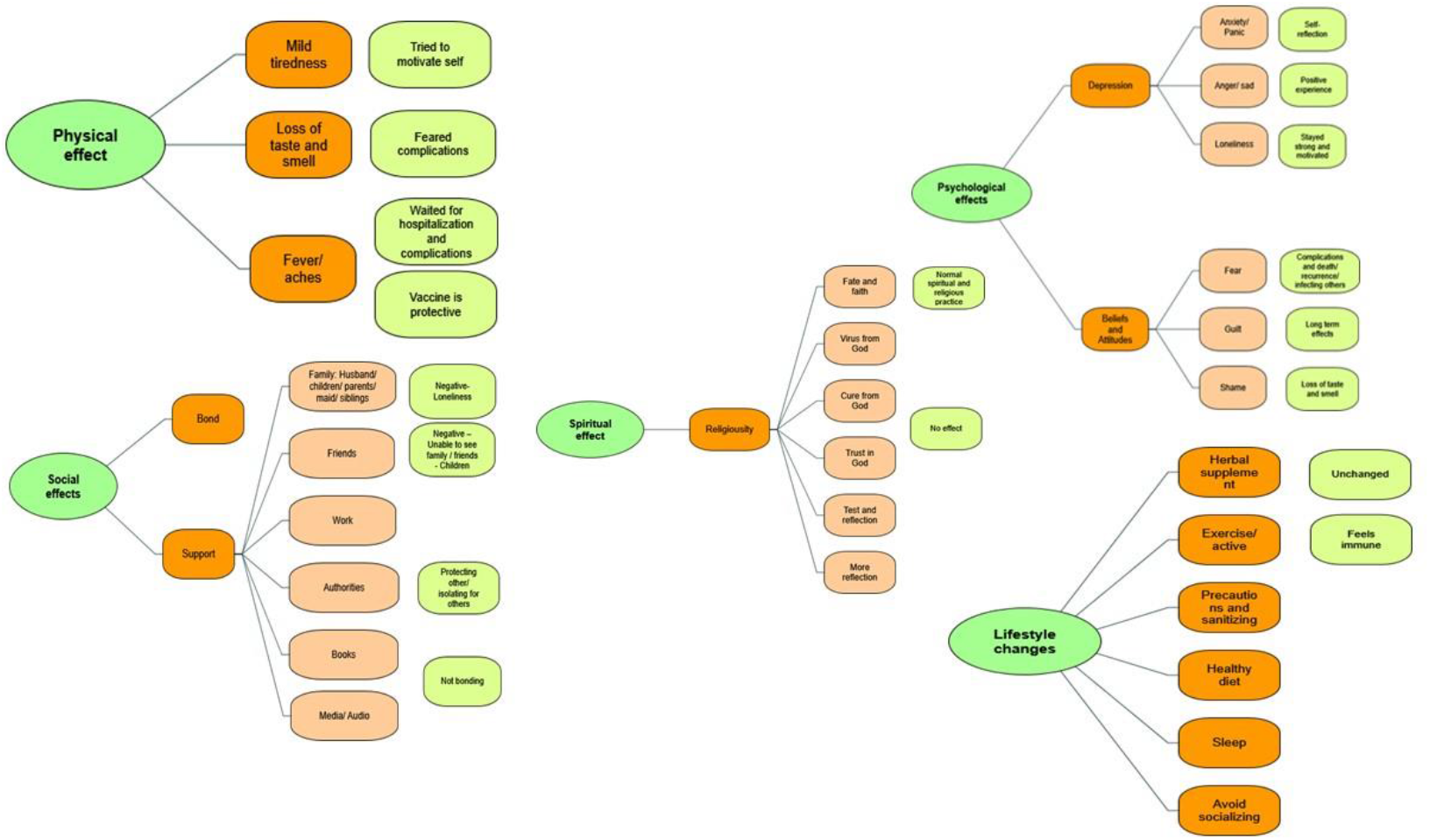
Themes tree

#### Description of the coding tree

The interview guide was divided into questions related to the physical, social, mental, lifestyle, and spiritual effects of a COVID-19 positive diagnosis on the participant. The coding was based on these five main effects explored.

#### Derivation themes

Themes derivation was based on the reported effects as described by participants (physical was based on what participants reported, social included the impact of direct family, friends, and social relations. Mental included the impact on psychological status during and post COVID-19 and returning to normal life. Lifestyle changes included impact on the use of a supplement, diet, sleep, and exercise. The last part was the impact on spiritual god, fear of death, and how that impacted on the beliefs and spiritual actions.

#### Software

All interviews were imported to NVivo 12 for analysis and coding. Thematic analysis using Braun & Larks 2006, six steps process was performed. The themes derived are presented in Figure 1, coded by main categories, and are presented according to their section by category. All effects were divided into the negative and the positive effect of the COVID-19 on the person’s status.

### Findings

Findings are classified based on the themes extracted from the interviews (Figure 1). Major themes are reported in Table 1, while identified and emerging themes are presented in Table 2. Major themes include the physical effects, social effects, psychological effects, spiritual effects, and lifestyle effects. Emerging themes include coping mechanisms, trust in authorities and health care system, appreciation of the role of the government, conspiracy theories, and media roles.

**Table 1:** Major themes reported by participants.

|  | **Positive** | **Negative** |
| --- | --- | --- |
| **Physical** | Acute: Moderate to mild able to cope with | Long term Effects |
| **Social** | Bond & Support | Loneliness |
| **Psychological** | Self-reflection/positive thinking/ appreciation of health | Anxiety/fear/anger/ stigma/shame |
| **Spiritual** | Strong relation with God | Unable to perform certain religious activities |
| **Lifestyle Effects** | Changed lifestyle to healthier die/exercise and sleep and more precautions | Feeling protected/ obsession with precautions |

**Table 2:** Emerging themes.

|  |  |
| --- | --- |
| **Coping** | • Self-reflection<br>• Time to stop and take a rest<br>• Catch up on missed chores<br>• Those who contracted COVID-109 at earlier times of the pandemic experienced more fear and anxiety because of uncertainties<br>• Blessing of being healthy |
| **Trust in government and Health Authorities and** | • Appreciation of the Effort<br>• We will be fine as we have all the support |
| **Conspiracy Theories** | • COVID-19 is man-made, vaccines are part of the conspiracy |
| **Education and Media** | • Media is creating Chaos, fear and anxiety<br>• More education is needed<br>• Media can be a great tool for education |

#### Physical effects

The majority of the participants had mild symptoms, from tiredness, and fatigue, loss of smell and taste, fever, and body aches. They have tried to motivate themselves, that even with their symptoms, they had to stay positive because they believe that their mindset can affect their physical well-being. Some feared more complications, and worsening of symptoms, awaiting hospitalization.

Some participants realized that the vaccine is protective, and have helped reduce the severity of the symptoms, especially the ones who took the two doses of the vaccine.

Most of our participants did not need hospital or experienced prolonged serious effects. Although so many have mentioned that they did not regain their normal smell of taste sensations. Some stated that the COVID-19 has impacted their general health and they have not regained their full strength. Three participants referred to some effects such as hair loss and irregular menses. Important to mention, some always fear losing their sense of smell, and taste, since it is a distinctive feature of COVID-19. Some participants mentioned that they were experiencing a bad smell even after they recovered.

#### Social Effect

The social effect of COVID-19 can be separated into positive and negative effects.

Positive effects include bond and support from family: husband/children/parents/maid/siblings, friends, work, health authorities, books, media/audio entertainment

Negative: loneliness, unable to see family, friends, children. Feeling of guilt unable to see parents or to look after them, stigma, stereotype, and feeling of shame and guilt. Protecting others/isolating others and no bonding.

All participants have received a text message and or a phone call, as soon as they turned out positive. Health authorities have shown tremendous efforts of helping and responding to those who turned out positive.

Participant 17 mentioned:

> “*Thank god I got COVID-19 here in the UAE, and not anywhere else, it is a blessing to be in the UAE, everything is vastly available, and the health authorities are just a call away*.*”*

In the UAE if a person is infected with COVID-19 with mild-moderate symptoms, they have the option to stay at home in isolation, or one of the isolation centers. The health authorities also provide food, laundry, and all necessities needed during isolation.

The majority of the participants have received increased support from their family members, husband, children, as well as co-workers. However, for participants that have no family members here in UAE, they have faced some difficulty having social support, nevertheless, with the use of social media, and the internet, the availability of video calls, they were able to keep in touch with loved ones.

Some took time to read books, watch YouTube videos, and keep themselves company with today’s technology to pass time, and bonded with themselves.

One participant mentioned the positive effect of an opportunity to look after her mom while admitted to the hospital for COVID-19 too. She finished her isolation time with her mom staying at the hospital to look after her. She expressed her blessed feeling for being positive at that time.

#### Psychological effects

The main negative psychological impact sub-themes extracted include Depression, anxiety/panic, anger sad, stigma, guilt, shame, fear of death, fear of complications, and recurrence.

Patients reported shock, anger, and anxiety as the majority of feelings on the discovery of their positive status. Most of the participants reported feeling sad, angry, shocked, and scared. Some reported that the anxiety continued throughout their isolation and felt anxious until they received were negative and finished the isolation. Their panic was either because of the COVID-19 status or because of the dilemma they faced on how they would break down the news to their families and colleagues from work. This feeling was reported mostly among the ones who got infected at the beginning of the pandemic, where uncertainty and fear of death were at their peak.

Participant 4 and her husband were infected with COVID-19 at the very beginning of the pandemic and she quotes:

> *“Fear and anxiety were at their peak at that time, since it was the first time to see a high morbidity, and mortality rates, as well as the information from the media, made it seem like it is the end of the world to get COVID-19”. She also mentions, that her husband was so scared of having COVID-19, that he wrote his ‘will’ because he believed he would die, from what he understood, and saw the start of the pandemic”*.

On a positive note, some participants found that being in isolation was for their benefit. They took some time to self-reflect, and time to take care of themselves, since they understood that we are living in a high-tech era, where most of the people are usually busy, working, with social obligations, and during their illness, taking time off they had the time to reflect and do the things they did not have time for before. Some were looking at the bright side, staying strong, and motivated, and that this illness will pass.

Participant 1 referred to this matter by saying:

> “*your mental health affects your immunity, and when you have a positive mindset towards the virus and illness, you can boost your immunity, and recover faster*.*”*

Some people indicated that when they lost their smell and taste senses, they felt the importance of appreciating these abilities by reflecting on what they had and what they lost.

Participant 28 said:

> “*When I got the COVID-19 I felt the blessings of tasting food and smelling*”

#### Beliefs and attitudes: fear, guilt, and shame

Almost all participants mentioned that they believe the virus is from God, and only God can end this pandemic, we have to pray and believe it is happening for our good. Also, the majority of the participants agreed that the COVID-19 virus is like any other virus, and it will require the contribution of the community, health authorities to fight the virus, and end the pandemic. Some participants were afraid of complications that are still unknown about the virus, fear of death, reoccurrence of the disease, and of course infecting others (peers from work, loved ones). Participant 12 mentions how a strong faith in God is needed in this pandemic, she quotes:

> “*Everything is from God, this is all already destined to happen, and we have no control of this, except to trust in God, and do what the authorities tell us to do, of course, we might fear future complications, but we need to keep a strong faith in God, and pray that this will all end*.*”*

Participant 14 said:

> “*The recovery period post corona is a new concept among families /People are afraid of corona patients although recovered”*.

Fear of losing one’s job is a major issue, especially how everything has changed since the pandemic has started. One of the participants has lost his job because of having COVID.

Fear of transmitting the disease to loved ones has resulted in avoidance of family gatherings, which may have led to feelings of hurt and shame increasing the burden of stigma. Stigma from family members and peers from work has been noticed towards those who are presumed to be affected, are undergoing treatment, or even ones who have recovered from the disease. Nowadays, the term “covid-positive” has become a stigma, and some might fear telling other family members, colleagues from work, about having COVID-19, due to numerous reasons: fear of isolation, being far from loved ones, being blamed for the illness, and for being careless.

Participant 9 expressed:

> “*Of course I felt stigma, especially with the people I see regularly, and of course I understand the paranoia, and how the transmission rates might scare people, but to stigmatize, especially my children, my neighbors would not let her children play with mine in the playground just because I had COVID, even though my kids are negative. There is definitely stigma, and it does not only affect the person who has the infection, but also the people living with that person can also feel stigmatized*.*”*

Another participant mentioned how their neighbors refrained from visiting them because of their fear to get COVID even after a while of their recovery and being negative.

#### Spiritual Effects

Most of the participants said that the experience either improved their spirituality and self-reflection or had no influence whilst none of them stated that the infection and the experience had any negative spiritual or religious effect.

Almost all participants agree that this is fate and faith. The virus is from God, and the cure is from God too. They trust God knows what is best, and it is a test for us. Some have increased their prayers during illness, while some stayed the same. Keeping a positive mindset and believing all is from God, have made it easier for them during isolation.

Participant 18 said:

> “*During any illness, you do get closer to Allah, and you keep making Dua, because, in the end, you have no one but Allah to talk to about how you are feeling, you know with Quran, Dua, and Praying”*.

Participant 12 said:

> “I *had more time during isolation to read Quraan on my own, do more prayers at night, and at that time I had the opportunity to be closer to Allah, I had more time to do the things I was really behind with my God, I used to finish Surat Al Baqara (a chapter in the Holy Quran) in one day, and I feel at that time, really my spirituality level were really high at that time, and this gave me more positive power and energy, that I will come out of this stronger than before inshaAllah (God willing)”*.

Participant 28 said:

> *“When I got the COVID-19 I felt the blessings of tasting food and smelling “*

#### Lifestyle Effects

Participants started paying attention to their mental health, and spending more time to rest, and relax during their illness, and keeping a positive mindset. Participants who had more physical symptoms reported that it was difficult to pray, move, and be active in the first few days of their illness. However, they tried their best to walk, move around, and eat better during isolation. After the COVID-19 infection, so many have stated that they returned to their normal lifestyle which they considered to be a healthy one. Few have stated that they decided to follow a healthier lifestyle after their COVID-19 experience to improve their immune system and to regain their full health.

Participant 17 mentioned:

> *“Ever since COVID happened, I made sure all we buy from the groceries are healthy, organic food, I limited white bread, and I increased their fruits and vegetable intake, to make sure that they have a healthy lifestyle to help them improve their immunity, for the whole family, also limit their time with watching TV, and being active at home*.*”*

Participants also mentioned that they were boosting their immune system through having vitamin C, zinc, honey, ginger, and black seed.

Since here in UAE, most people believe in the healing properties of herbal supplements. Almost all of the participants have taken herbal supplements, and recipes from families and friends, that claim to relieve their symptoms. Some participants have tried to stay active, and exercise even after being recovered to help protect them from future illness, and reoccurrence.

Some participants who have had the vaccine, and got covid, have reduced their precautions, and sanitizing, still wearing a mask, but less sanitizing. While some, still sanitize, more or so as the beginning of the pandemic.

Participant 11 expressed:

> “*We wear masks because it is mandatory, but I believe when I took the vaccine, and I got COVID-19, I am now more immune, and no need for sanitizing all the time*.*” While others mentioned, “Since I work in the hospital, I still do all the precautions, even though I am vaccinated, and I got infected with COVID-19, my family and I still sanitize everything, and taking more or so the same precautions as before*.*”*

A few of the participants have reduced their family gatherings and still avoid socializing with family members especially the ones who have co-morbidities.

Some stated that they have an unchanged lifestyle as they have had a healthy lifestyle in general before the COVID-19 while others feel immune especially after getting the vaccine and having a positive COVID-19 infection.

Participant 14 said:

> *“As you know, my mother and father have many co-morbid conditions, that even my husband told me to wait, we should not be in a hurry and go visit everyone as soon as we are negative, because we are not sure if we can still transmit the disease or not? It is better to be careful than sorry*.*”*

On another note, participant 11 said:

> *“My life has not changed since I recovered from COVID-19, I still visit my family, and have gatherings, I even rewarded myself after I became negative twice to a trip to the Maldives, I am not doing the precautions as before, but I still listen to the authorities, and wear my mask of course*.*”*

### Emerging Themes

#### Coping

Participants described utilizing different and various coping mechanisms. Some have tried self-motivation, convincing themselves that their situation is temporary. Others resorted to reading books, Quran, watching online videos including motivational, and self-help videos, to help cope with the illness.

Participant 12 saod:

> “*Here is where the isolation was a blessing in disguise, for the first time in a very long time, I had time to read my books, watch videos, and documentaries, to entertain myself, it helped me cope with the situation, and I understood that it’s just going to be for a while and that I can do all the things I could not do, because of life’s high demanding work, family obligations. The isolation helped me, reflect, and do the things I enjoy the most*.”

Few participants n.5 (including 3 working in the medical field) have taken isolation as an opportunity to rest, to self-care, and to catch up with their life. The majority of the participants have used social networking through zoom, phone, and social media to cope with isolation. Some focused on having a healthy diet, and a good diet to help with their immunity. Finally, religious activities such as praying and reading Quran was commonly used by most participants

#### Trust in government and health authorities

One of the very interesting emerging themes was the trust in the health care system and the government’s role in managing the pandemic. Many of the participants expressed their trust in the treatment protocols and management of the positive cases, indicating that it had a great impact on their journey with the COVID-19 psychologically, physically, and socially. Although some had some negative, comments and showed concerns that in Dubai there was not much support such as the one provided in Abu Dhabi, however, overall many had full trust and appreciation.

#### Conspiracy theories

When asked about the COVID-19 and their experience around 25% of the participants indicated that COVID-19 is a man-made virus aimed to destroy communities. For others, vaccination was not an option due to their disbelief in the whole situation. Even though some had their doubts, they were still willing to deal with them.

Participant 17 mentioned

> *“Even if it is a conspiracy we have to accept it and face it”*

For some the doubt existed in the vaccine, Couple of participants expressed their concerns that once they took the vaccine they became positive.

> Participant 18 said
>
> *“I believe that it was the vaccine that has infected me with COVID-19”*

#### Education and media

Finally, it is highly important to mention that the time of infection and the duration played a role on the impact of people psychological and lifestyle as those who were infected earlier on during the pandemic had lower knowledge and were receiving confusing messages and information from authorities, health care professionals and media.

Participant 8 said:

> “*The last year experience and people stories of recovery make it easier and not scary,”*

Participant 17 also said:

> *“Number of deaths and uncertainties in the earlier time (2020) was scary”*

Some have commented on the role of media in exaggerating news and causing chaos calling for more restrictions and actions from authorities,

## Discussion

The study explored the physical, social, psychological, spiritual, and lifestyle effects of a positive COVID-19 diagnosis on a sample of the UAE population. Physical effects reported by participants varied from loss of smell and taste to tiredness, fatigue, Flu-like symptoms, and shortness of breath. Some social effects were reported as positive effects such as bonding with the family, appreciating family, friends, and neighbors’ support. The negative social effects of a COVID-19 infection included stigma of a positive case, displaying fear of the persons with COVID-19 even after recovery, being away for at least 14 days from loved ones, fear for loved ones, losing on a job, and financial loss as well as being away from friends and daily activities. Psychological and mental effects also can be divided into negative and positive. The positive psychological effects of being infected by COVID-19 were feeling blessed and taking time for self-reflection and meditation. The infection served for so many as an opportunity to catch up with missed activities such as reading, watching videos, and connecting with others through social media and phones. However, the negative effects appeared to be more as fear, anxiety, anger, and distress were reported by most participants. Many reasons were recalled by participants such as the fear of death, complications, loss of a loved one, and isolation. Spiritual effects for most were also positive as they claimed that the COVID-19 made them closer to God and helped them on improving their spiritual practices. Although for some who were infected during Ramadan, the COVID-19 was considered to be a barrier to practicing their religious duties. Finally, so many reported thinking more about a healthier lifestyle to strengthen their immunity and to avoid getting sick again. Many participants mentioned they will get the vaccine and using vitamins, supplements, honey, herbal remedies during the period of isolation.

Emerging themes extracted include believing in conspiracy, trust in government, the role of health care, and the health system.

The UAE health system has proven to be one of the efficient during the COVID-19 situation with vaccination strategies and mass testing being effective in curbing the spread of the virus and reducing hospitalizations and severe complications (Al-Hosani et al. 2021; Suliman et al. 2021). However, there is always a need to improve the social and health support for patients with COVID-19 and their close contacts. Many participants reported being very pleased with the health authorities responded to the pandemic in terms of the health services provided (care and attention received). This is specific for residents in Al Ain and Abu Dhabi, in the emirate of Abu Dhabi. The Department of Health Abu Dhabi has been Residents from other emirates reported not so positive feedback from the support received from their respected health authorities. These differences might be due to the different health authorities and regulatory bodies of each emirate, the UAE health system is unique comprising federal and Emirati level entities. During the pandemic, a coordinated response from the Ministry of Health and Prevention, the Department of Health Abu Dhabi, the Abu Dhabi Public Health Center, and the Dubai Health Authority activated a National Technical Advisory Committee to coordinate different health policies and actions across different emirates. The *National Guidelines for Clinical Management and Treatment of COVID-19* were issued in response to global evidence regarding the containment of the virus and treatment of the disease (National Clinical Committee for COVID-19 Management 2020) and each regulatory authority issued its guidelines and coordinated the responses within their health systems.

The findings of this study highlight the importance of close support during the infection and after immediate recovery and post-recovery. Social and psychological support are highly needed especially to deal with the long-term effect of the social and psychological impact of the infection. Serafini et al (2020) reviewed the literature and found that resilience, loneliness, anxiety, loss of freedom, and wellbeing were the main psychological effects of COVID-19 on communities (Serafini et al. 2020). This may typically increase with being affected directly or having a loved one affected by the infection. The same authors and others discuss the impact of the COVID-19 pandemic on lifestyle and social isolation (Banerjee and Rai 2020; Park et al. 2021; S et al. 2020; Serafini et al. 2020). Study participants reported lots of changes during the infection with their lifestyle especially isolation, loss of work, and being away from others but other changes may last after the infection such as the increase in the use of precautions measures (handwashing, sanitizers, changes in diet and other lifestyle factors). Proper advice and education are mandatory in these cases, one of the participants was a psychologist who pointed out the increase in obsessive-compulsive behaviors among people due to the fear of infection. This was also questioned by Jelinek et al (Jelinek et al. 2021).

We noted during the interviews, that beliefs towards the virus have changed from people who got infected in 2020 and 2021. In 2021, people’s knowledge and acceptance of vaccination, fear of dying from COVID-19, and the unknown long-term and short-term effects have decreased with time and with more education. Studies assessing knowledge and awareness about COVID-19 and its vaccines have shown that positive attitudes have increased due to local governments’ efforts and global education (Lutfi et al. 2021).

A significant finding in this study was that those who had a positive infection end toward the end of 2020 and in 2021 described having less negative emotions and better psychological resilience. They referred to the fact that they had better knowledge about the COVID-19 effects, availability of the vaccines, and governments’ efforts to ease the pandemic. Although some participants expressed their satisfaction with the support they received, a big number of participants (almost 90%) reported their strong appreciation to the UAE government and health authorities reflected on satisfactory knowledge and favorable practice, with an overall high positive attitude (Lutfi et al. 2021). This finding is a reflection of the sentiments of the Emirati population and people residing in the UAE and.

The COVID-19 hotline provided by the health authorities identifies people who may be infected and does not focus on providing psychological support to help with the patients’ fears and doubts. Although improvement knowledge, awareness, and self-copying strategies are still critical in this pandemic.

It is crucial to educate newly diagnosed patients about COVID-19, and the most recent knowledge about it, to prevent people being overwhelmed with stigma from their society and the media. Another important point is to inform diagnosed patients with COVID-19, to report any mental or social issues, that can be solved with the course of their physical symptoms. Most of these patients have had all the support they needed, but we cannot deny that there is the need to focus more on mental and social well-being of these patients, because they understand that their immune system is affected by their mental health. The participants have perceived a very good support in terms of their physical health from the government and health authorities.

## Limitations

This is a qualitative study and we cannot generalize the results to the general population. However, being an explorative study further studies can build on our findings. Another limitation of this study is the COVID-19 situation, where lockdowns and the chaos caused in the society limited our access to participants and to conduct face-to-face interviews. Moreover, many participants approached were reluctant to discuss their experiences.

## Conclusion

The findings of this study indicate that people diagnosed with COVID-19 have perceived a very good support in terms of their physical health from the government and health authorities, but require social, psychological, and educational support during the infection period and post-recovery. This support may be needed for longer period to prevent further implications on individuals and families’ physical and psychological health.

## Data Availability

All data can be requested from the corresponding author

## Declarations

### Funding

The authors did not receive support from any organization for the submitted work.

### Conflict of Interest

The authors have no conflicts of interest to declare.

### Availability of data and material

Upon request to the corresponding author.

### Code availability

Not available.

### Authors’ contributions

IE conceptualized the study. IE, BH, MAL & NAL conducted the interviews and analyzed the data. IE, BH & MAL agreed on the themes. IE, MAL, MSP & NAL wrote and reviewed the manuscript. All authors accepted the manuscript for publication.

### Ethics approval

This study was performed in line with the principles of the Declaration of Helsinki. Approval was granted by the Social Sciences Ethics Committee of the United Arab Emirates University (ERS_2021_7264).

### Consent to participate

Informed consent was obtained from all individual participants included in the study.

### Consent for publication

Authors are responsible for correctness of the statements provided in the manuscript.

## Acknowledgements

The authors would like to acknowledge all participants and the Emirates Centre for Happiness Research at the United Arab Emirates University for their support on this research.

## Notes

### Competing Interest Statement

The authors have declared no competing interest.

### Funding Statement

No external funding was received

### Author Declarations

Social Sciences Ethics Committee of the United Arab Emirates University (ERS_2021_7264).

